# The effects of physiotherapy programs in COVID-19 patients during admission in the hospital

**DOI:** 10.1101/2023.01.27.23285094

**Authors:** Netchanok Jianramas, Veeranoot Nissapatorn, Chaisith Sivakorn, Maria de Lourdes Pereira, Anuttra (Chaovavanich) Ratnarathon, Chenpak Salesingh, Eittipad Jaiyen, Salinee Chaiyakul, Nitita Piya-amornphan, Thanaporn Semphuet, Thanrada Thiangtham, Kornchanok Boontam, Khomkrip Longlalerng

## Abstract

**Background and aims:** Several recommendations exist regarding the role of physiotherapy programs (PTPs) in COVID-19 patients. However, none of the studies examines the frequency of bedside PTPs during admission. Thus, this study aimed to compare the different bedside PTPs frequencies on the survival rate, length of hospitalization (LoH), referrals to the intensive care unit (ICU), and in-hospital complications. The safety of patients and the physiotherapist was also investigated.

**Methods:** Fifty-two COVID-19 patients were equally assigned into two groups matched on gender and age (1:1 ratio). Experimental group one received 1-2 times of PTPs during hospitalization, and experimental group two received daily PTPs until hospital discharge. The primary outcomes were the survival rate, LoH, referrals to ICU, and in-hospital complications. The secondary outcomes were the adverse events for patients and the number of physiotherapists who contracted with COVID-19.

**Results:** Most participants were classified as having mild to moderate COVID-19 with a mean age of 45 years. There were no differences between groups in all primary outcome measures (all p > 0.05). The overall survival rate was 98%. One participant from the Ex-G2 group was referred to the ICU. Two Ex-G1 and four Ex-G2 participants had complications. There were no immediate serious adverse events found after PTPs for both groups. None of the physiotherapists tested positive for COVID-19.

**Conclusion:** In COVID-19 patients with mild to moderate conditions, one to two bedside PTPs were enough to achieve the same results as patients who received daily PTPs. PTPs were safe for COVID-19 patients, and physiotherapists.

**CLINICAL REGISTRATION NUMBER:** Thai Clinical Trials, https://www.thaiclinicaltrials.org/, TCTR20210823004.

## Introduction

Since 2019, the human coronavirus disease 2019 (COVID-19) has spread throughout the world, including Thailand [1, 2]. This emerging disease directly affects the patient’s respiratory system [3]. Its clinical signs and symptoms range from fever, cough, chill, short, shallow, and difficult breathing, fatigue, malaise, headache, anosmia, ageusia, sore throat, stuffy nose and runny nose, nausea and vomiting, and diarrhea [3-5]. Patients with more severe conditions experience more pronounced signs and symptoms, which are caused by pneumonia [6]. Physiotherapy programs (PTPs) are commonly suggested for adults with pneumonia who are intubated and mechanically ventilated, promoting clearance of secretions and lung compliance [7]. An expert physiotherapist suggested that PTPs should be applied to COVID-19 patients if there is an indication of pneumonia without exudate consolidation, mucous hypersecretion and difficulty clearing secretions, functional decline, and (at risk of) intensive care unit (ICU)-acquired weakness [8-10]. PTPs range from prone positioning, postural drainage, breathing exercises and devices (e.g. positive expiratory pressure and inspiratory muscle training), ventilator settings, positioning, as well as functional training, exercise, and early mobilization [9, 11-13].

Notably, suggestions of most experts and some of the studies agreed that patients with severe symptoms or after recovery from the intensive care unit derive benefits from PTPs [9, 11 - 14]. Conversely, some researchers have suggested that PTPs be contraindicated during the acute phase because the acute effects of the exercise program may cause an increase in pro-inflammatory cytokines and viral replication [15, 16]. Nevertheless, a recent study has shown an improvement in immune function after two weeks of moderate aerobic exercise,[17] which is supported by previous review studies [18, 19]. Interestingly, most previous studies have investigated the effects of PTPs in the acute phase or sub-acute phase in severe to critically ill patients with COVID-19 [11, 12, 14, 20, 21]. However, there are limited studies on the lesser severity of COVID-19 patients. Some of the recommendations agreed that conventional PTPs and mild to moderate physical exercise can be applied to COVID-19 patients with mild to moderate severity [13].

To our knowledge, bedside PTPs should be applied to patients as soon as and as often as possible. Some studies have discovered that more frequent bedside PTPs are more effective at lowering mortality rates, hospitalization days, and respiratory infections in ICU patients [22]. However, little is known about the effects of different bedside PTP frequencies in COVID-19 patients. Thus, this study aimed to compare the different bedside PTP frequencies on the survival rate, length of hospitalization (LoH), referrals to the ICU, and in-hospital complications. In addition, the safety of patients during and after performing PTPs was investigated. We hypothesized that COVID-19 patients receiving daily bedside PTPs would have a significantly higher survival rate, lower LoH, as well as fewer complications compared to those receiving fewer bedside PTPs. In addition, none of the COVID-19 patients had serious adverse events during and after PTPs, and no physiotherapists tested positive for COVID-19 infection.

## Methods

### Study design

A prospective, quasi-experimental study design was used to determine the effects of PTPs in the acute phase of patients with COVID-19. This study was conducted at Bamrasnaradura Infectious Diseases Institute, Nonthaburi Province, Thailand, from November 2021 to January 2022. The technical approval was granted by the Human Research Ethics Committee of the Bamrasnaradura Infectious Disease Institute, Nonthaburi Province (S020h/64), and the Human Research Ethics Committee of Walailak University, Nakhon Si Thammarat Province (WU- EC- AL- 3- 186- 64). Written informed consent was acquired from all participants. All procedures performed in this study involving humans were in accordance with the ethical standards of the institutional and/or national research committee and the 1964 Helsinki Declaration and its later amendment or comparable ethical standards.

### Study population

Participants were assigned into two groups and stratified by age and gender. General inclusion criteria included adults aged 18-65 years old with 1) a nucleic acid test-confirmed diagnosis of SAR CoV-2 infection, 2) hospitalization due to any clinical signs and symptoms of pneumonia, 3) no communication problems, and 4) ability to use an online mobile phone application. For specific inclusion criteria, patients were recruited if one of the following criteria was indicated: [8, 9] 1) COVID-19 with risk factors for severe disease, 2) confirmed case of pneumonia with hypoxia (resting blood oxygen saturation (SpO_2_) < 96% or the presence of exercise-induced hypoxemia defined as a reduction in SpO_2_ > 3% compared to baseline, 3) inability to expel secretions caused by prolonged immobilization and respiratory muscle weakness, 4) presence of breathlessness or dyspnea needing oxygen therapy (presence of signs of pneumonia with lung consolidation by chest radiograph or computerized tomography or lung ultrasound), and 5) presence of functional limitation caused by prolonged hospitalization or prolonged ICU stay or prolonged use of a respirator or oxygen device. Exclusion criteria included 1) unwillingness or inability to follow the study protocol and 2) active participation in another study.

### Characteristics and classification of disease severity of COVID-19

COVID- 19 patients were categorized as mild, moderate, severe, and critical according to the World Health Organization definition [23]. Mild severity (mild pneumonia) was defined as a person with the following symptoms: fever, cough, fatigue, anorexia, shortness of breath, and myalgia. Moderate severity (moderate pneumonia) was defined as a person with more pronounced signs and symptoms of viral pneumonia or hypoxia. Severe COVID-19 was defined as a person with pneumonia accompanied by any signs and symptoms, including respiratory rate > 30 times/min, and SpO_2_ < 90% on room air. Critical disease was defined as a patient with ARDS [23]. In addition, a laboratory investigation including the open reading frame gene1ab (ORF1ab) and the envelope (E) gene of SARS-CoV-2, complete blood count (CBC), kidney and liver function, tissue damage markers (lactate dehydrogenase (LDH)), inflammatory markers (i.e. erythrocyte sedimentation rate (ESR) and C-reactive protein (CRP)), chest radiography, and medications were used to classify the severity of patients at baseline. A patient who has been administered a combination of antiviral drugs and/or received corticosteroid treatment was classified as a moderate to severe case.

### Intervention (physiotherapy programs (PTPs)

Participants in Experimental Group One (Ex-G1) received only a one-time bedside PTPs for the first few days after hospital admission. However, some patients remained confused of the program, and thus a second bedside PTPs was taught to patients. Meanwhile, participants in Experimental Group Two (Ex-G2) had daily bedside PTPs until hospital discharge. PTPs comprised of breathing exercises, secretion removal techniques (coughing, huffing, and positioning), active chest trunk mobilization, active exercise of both limbs, and early progressive mobilizations/ ambulation [9, 11-13]. The BreatheMax®V.2 device (C&D Biomedical, Thailand) that operates based on a principle of positive expiratory pressure (PEP) and oscillating incentive spirometer (OIS) was given to Ex-G2 patients. In addition, all patients were asked to join a closed private group via a mobile application platform. Some of the patients were encouraged and monitored their signs and symptoms directly on their private mobile application or ward phone, especially Ex-G1 participants who were taught only one-time PTPs.

### Safety considerations for COVID-19 patients in assigning the physiotherapy programs

Prior to perform bedside PTPs, all patient data, vital signs, and medical records were intensively reviewed by the physiotherapists. Patients were questioned on their current signs and symptoms. Permission was not granted for physiotherapist treatment if the patient had any of the following regardless of whether they were on a ventilator: fraction of inspired oxygen (FiO_2_) > 0. 6, SpO_2_ < 90%, respiratory rate (RR) > 40 times/ min, positive end-expiratory pressure (PEEP) > 10 cmH_2_O, ventilator resistance, unstable cardiovascular signs (systolic blood pressure > 180 mmHg or diastolic blood pressure > 110 mmHg, heart rate < 40 or > 120 beats/min) [24]. If SpO_2_ decreased > 3% from baseline during the PTPs session, they were not allowed to continue the session.

### Safety for physiotherapists to prevent SARS-CoV-2 transmission

Physiotherapists were trained to use personal protective equipment (PPE) properly before performing bedside PTPs [9]. Importantly, all physiotherapists had to strictly follow aerosol, airborne, and contact precautions [9]. For physiotherapist safety considerations, all of them must have 1) a minimum of two doses of the COVID-19 vaccine before initiation of the study for at least 2 weeks, 2) good health with no comorbidities, 3) age of < 45 years [9], and 4) at least two years of experience in chest physiotherapy. All physiotherapists were tested weekly for COVID-19 infection using reverse transcription-polymerase chain reaction (RT-PCR), and rapid antigen test kit (ATK).

### Outcome measures

The primary outcomes were survival rate, length of hospital stay (LoH), number of patients who were referred to ICU, and rates/types of complications. The secondary outcome was the safety of the patients, which was indicated by the minor and serious adverse events during and after each physical therapy session. A minor adverse event was defined as an event that slightly affected the patient, requiring time for rest, and recovery within 15 minutes for example; dizziness, nausea and vomiting, postural hypotension (blood pressure drop > 10 mmHg from baseline), fatigue, and SpO_2_ drop greater than 3% from baseline. A serious adverse event was defined as an event that needed urgent assistance, for example, medications or resuscitation. In addition, the other secondary outcome was the safety of the physiotherapists. The physiotherapists were tested for COVID-19 infection using RT-PCR and ATK every week.

### Laboratory tests

Blood samples were collected from all participants in the antebrachial area. RT-PCR was performed using a Cobas® 6800 SARS-CoV-2 assay on the Cobas ® 6800 platforms (Roche Diagnostics, Basel, Switzerland) to identify the presence of the ORF1ab and the E-gene of SARS-CoV-2. ATK was tested using a Singclean (Hangzhou Singclean Medical Products, China). CBC was examined using an electrical impedance (Mindray Model CAL 6000, China). Meanwhile, the renal function test, liver function test, and LDH were examined using photometry (Beckman Coulter, USA). ESR was examined using the Westergren method (Mini-VES, Italy). CRP was examined using immunofluorescence (UNICELL-S, China).

### Statistical analysis

Descriptive data was reported as mean±SD or median (IQR) if it indicated normal or non-normal distribution of data, respectively. The sample size was calculated using the G-Power program Version 3.10. The partial eta square (η^2^) was set at a medium level. Assuming a moderate effect size of 0. 25, a power of 90%, and a dropout rate of 20%, thus we planned for enrolling 52 participants. The parametric distribution was assessed using the Shapiro-Wilk test. Nominal data were summarized as frequencies and percentages. Categorical variables were compared using the Chi-squared test or Fisher’ s exact test, as appropriate. Continuous variables were compared by Independent t-test or Mann-Whitney U test, as appropriate. Data were analyzed using the Statistical Package for Social Sciences Version 22.0 (IBM Inc., Armonk, NY). The level of significance was taken at 0.05 or 5%.

## Results

### Participant characteristics

Sixty-six COVID-19 patients were identified as eligible for participation. Fourteen participants were excluded as shown in Fig 1. Most participants were classified as having the Delta variant (one participant had the Omicron variant). The mean age of all participants was 45±14 years, and 46% were men. Most participants (87%) were vaccinated against COVID-19. The COVID-19 severity was similar between groups (Table 1 and Table 2). Most participants (n = 49, 94%) were categorized as non-severe cases, which ranged from mild to moderate conditions of COVID-19. The proportions of disease severity were mild (n = 26; 50%), moderate (n = 23; 44%), and severe (n = 3; 6%) conditions. Most participants had comorbidities (n = 41, 79%). At the first bedside physiotherapy visit, three Ex-G1 and twelve Ex-G2 patients were admitted to the intensive care unit (ICU). None of the baseline characteristics were significantly different between groups (all p > 0.05) (Table 1 and Table 2). Most hematologic parameters were in the normal range, except for the inflammatory markers of ESR and CRP (Table 2).

**Table 1.** Comparison of baseline characteristics between groups.

|  | **Ex-G1 (n = 26)** | **Ex-G2 (n = 26)** | **P Value** |
| --- | --- | --- | --- |
| Men | 12 (46%) | 12 (46%) | 1.000 |
| Age, mean (SD), years | 43.50 (13.34) | 45.92 (15.24) | 0.545 |
| Age > 60 y | 4 (16%) | 5 (19%) | 0.714 |
| Education level |  |  |  |
| Higher than bachelor degree | 4 (16%) | 1 (4%) | 0.168 |
| Bachelor degree | 9 (35%) | 7 (27%) |  |
| Secondary school | 4 (16%) | 10 (38%) |  |
| Primary school | 7 (27%) | 8 (31%) |  |
| No formal education | 2 (8%) | 0 |  |
| On oxygen devices |  |  |  |
| Nasal cannula | 1 (4%) | 3 (12%) | 0.350 |
| High flow nasal cannula | 0 | 1 (4%) |  |
| Mechanical ventilator | 0 | 0 |  |
| Comorbidities |  |  |  |
| No | 4 (15%) | 7 (27%) | 0.308 |
| Yes<sup>a</sup> | 22 (85%) | 19 (73%) |  |
| BW, median (IQR), kg | 69.50 (57.00, 89.25) | 68.00 (57.00, 85.00) | 0.905 |
| BMI, median (IQR), kg/m<sup>2</sup> | 25.06 (22.83, 32.34) | 26.88 (23.18, 30.99) | 0.504 |
| Classification of BMI<sup>b</sup> |  |  |  |
| Obese | 13 (50%) | 16 (62%) | 0.606 |
| Overweight | 6 (23%) | 5 (19%) |  |
| Normal | 7 (27%) | 5 (19%) |  |
| COVID-19 disease severity |  |  |  |
| Mild | 15 (58%) | 11 (42%) | 0.538 |
| Moderate | 10 (38%) | 13 (50%) |  |
| Severe | 1 (4%) | 2 (8%) |  |
| Duration from admission to initiating PT, mean (SD), days | 1.62 (0.80) | 1.69 (1.09) | 0.773 |
| Antiviral medication use |  |  |  |
| Favipiravir | 24 (92%) | 22 (100%) | 1.000 |
| Mixed antiviral drug | 13 (50%) | 10 (40%) | 0.402 |
| Corticosteroid drug | 3 (12%) | 8 (32%) | 0.090 |

Data expressed as n (%) unless otherwise stated.
<sup>a</sup> Comorbidities included hypertension, diabetes mellitus, cerebrovascular accident, thyroid, allergy, glucose-6-phosphate dehydrogenase deficiency, chronic hepatitis B infection, thalassemia, systemic lupus erythematosus, enlarged prostate, gout, old tuberculosis infection, migraine, and chronic kidney disease.
<sup>b</sup> Obese = BMI ≥ 25; overweight = $23 \leq$ BMI < 25; and normal = $18.5 \leq$ BMI < 23.
Abbreviation: BMI, body mass index; BW, body weight; COVID-19, Coronavirus Disease 2019; ICU, intensive care unit; IQR, interquartile range; PT, physiotherapy; SD, standard deviation

**Table 2.** Comparison of baseline blood biomarkers between groups.

|  | Ex-G1 (n = 26) | Ex-G2 (n = 25<sup>a</sup>) | *P* Value |
| --- | --- | --- | --- |
| ORF1ab gene, Ct | 20.81 (17.76, 23.64) | 20.56 (18.59, 24.10) | 0.859 |
| E-gene, Ct | 21.70 (18.09, 24.09) | 21.07 (19.03, 23.93) | 0.861 |
| Complete blood count |  |  |  |
| WBC, mean (SD), $\times 10^3/\mu\text{L}$ | 6.90 (1.74) | 7.01 (2.44) | 0.856 |
| RBC, mean (SD), $10^6/\mu\text{L}$ | 5.07 (0.61) | 4.83 (0.64) | 0.167 |
| HGB, mean (SD), g/dL | 13.27 (1.56) | 13.4 (1.45) | 0.661 |
| HCT, mean (SD), % | 40.62 (4.44) | 40.84 (4.01) | 0.850 |
| RDW, % | 13.90 (13.00, 14.53) | 13.50 (12.60, 13.90) | 0.086 |
| Platelet, $\times 10^3/\mu\text{L}$ | 254.00 (215.50, 293.75) | 254.00 (207.50, 332.00) | 0.706 |
| Neutrophil, mean (SD), % | 60.77 (13.39) | 63.56 (12.94) | 0.453 |
| Lymphocyte, mean (SD), % | 28.46 (12.34) | 26.84 (12.83) | 0.647 |
| Monocyte, % | 7.50 (6.00, 10.00) | 6.00 (5.00, 9.50) | 0.209 |
| Eosinophil, % | 1.00 (0.00, 3.00) | 1.00 (0.00, 2.00) | 0.486 |
| Basophil, % | 0.00 (0.00, 1.00) | 0.00 (0.00, 1.00) | 0.925 |
| Kidney function test |  |  |  |
| BUN, mg/dL | 12.50 (9.00, 15.00) | 11.00 (8.00, 17.00) | 0.769 |
| Creatinine, mg/dL | 0.74 (0.64, 0.88) | 0.71 (0.58, 1.04) | 0.850 |
| eGFR, mL/min/1.73m<sup>2</sup> | 106.01 (87.45, 115.83) | 107.06 (71.72, 122.14) | 0.880 |
| eGFR stage, n (%) |  |  | 0.086 |
| Stage I | 19 (73%) | 17 (68%) |  |
| Stage II | 7 (27%) | 4 (16%) |  |
| Stage III | 0 | 4 (16%) |  |
| Liver function test |  |  |  |
| Total protein, mean (SD), g/dl | 7.74 (0.75) | 7.63 (0.58) | 0.655 |
| Total bilirubin, mean (SD), mg/dl | 0.49 (0.18) | 0.52 (0.17) | 0.855 |
| Alkaline phosphate, U/L | 69.00 (55.00, 88.00) | 67.00 (52.00, 78.50) | 0.516 |
| AST/SGOT, U/L | 26.00 (22.50, 36.50) | 26.00 (23.00, 30.00) | 0.437 |
| ALT/SGPT, U/L | 24.00 (17.00, 47.50) | 22.00 (14.00, 39.00) | 0.166 |
| LDH, U/L | 189.30 (159.50, 207.15) | 176.30 (163.55, 242.85) | 0.777 |
| Inflammation biomarkers |  |  |  |
| ESR, mean (SD), mm/hr | 26.92 (13.06) | 31.12 (19.00) | 0.297 |
| CRP, mg/dL | 8.26 (3.56, 17.94) | 13.89 (5.80, 25.66) | 0.122 |

Data expressed as median (IQR) unless otherwise stated.
<sup>a</sup> n = 25 due to a laboratory test of one participant that the clinician did not order.
*Abbreviation:* ALT/ SGPT, alanine aminotransferase/ serum glutamic-pyruvic transaminase; AST/ SGOT, aspartate aminotransferase/ serum glutamic-oxaloacetic transaminase; BUN, blood urine nitrogen; CRP, C-reactive protein; Ct, threshold cycle; E-gene, envelope gene; eGFR, estimated glomerular filtration rate; ESR, erythrocyte sedimentation rate; HCT, hematocrit; HGB, hemoglobin; LDH, lactate dehydrogenase; ORF1ab-gene, open reading frames ab gene; RBC, red blood cell; RDW, red blood cell distribution width; WBC, white blood cell

**Fig 1.** Flow diagram for participants throughout this study.

### Primary outcomes

There were no significant differences in all outcome measures found between the intervention groups (p > 0. 05) (Table 3). The survival rate of all COVID-19 patients was 98% in this study. The survival rate of COVID-19 patients with mild and moderate conditions was equal (100%) between the groups. There was only one Ex-G2 participant referred to the ICU room after enrollment to the study because of influenza. There were two Ex-G1 and four Ex-G2 participants with complications after receiving PTPs (Table 3).

**Table 3.** Outcome measurement comparison between groups.

|  | Ex-G1 (n = 26) | Ex-G2 (n = 26) | *P* Value |
| --- | --- | --- | --- |
| Survival | 26 (100%) | 25 (96%) | 1.000 |
| Death | 0 | 1 (4%) | 1.000 |
| LoH, median (IQR), days | 10.00 (9.00, 11.80) | 10.00 (10.00, 12.00) | 0.117 |
| Patients referred to ICU | 0 | 1 (4%) | 0.313 |
| Complications |  |  |  |
| Influenza | 0 | 1 (4%) | 0.555 |
| Bacterial infection | 2 (8%) | 2 (8%) |  |
| Cardiac arrest | 0 | 1 (4%) |  |
| None | 24 (92%) | 22 (84%) |  |
| Minor adverse event |  |  |  |
| Drop of SpO<sub>2</sub> > 3% from baseline | 0 | 8 (31%) | 0.018 |
| Dizziness | 1 (4%) | 1 (4%) |  |
| Nausea & vomiting | 2 (8%) | 0 |  |
| <sup>a</sup>Dyspnea | 2 (8%) | 1 (4%) |  |
| <sup>a</sup>Dyspnea & drop of SpO<sub>2</sub> > 3% from baseline | 0 | 1 (4%) |  |
| None | 21 (80%) | 15 (57%) |  |
| Average of PT bedside, median (IQR) | 2.00 (1.00, 2.00) | 6.00 (5.00, 7.00) | <0.001 |
| Number of patients receiving each PT program |  |  |  |
| Breathing exercise, cough/ huff training, active chest trunk mobilization, positioning, active exercise of UE and LE | 26 (100%) | 26 (100%) | 1.000 |
| Positive expiratory pressure devices | 0 | 26 (100%) | <0.001 |
| Out-of-bed exercise | 0 | 26 (100%) | <0.001 |

Data expressed as n (%) unless otherwise stated.
<sup>a</sup> Dyspnea caused by continued cough during breathing exercise
Abbreviation: ICU; intensive care unit; IQR, interquartile range; LE, lower extremity; LoH, length of hospital stay; PT, physiotherapy; UE, upper extremity

### Secondary outcomes

Five Ex-G1 and eleven Ex-G2 COVID-19 patients had minor adverse events during and after the PTPs (Table 3). Among eight Ex-G2 participants, there were 14 sessions (9%) of the bedside PTPs that had a SpO_2_ drop > 3% from baseline (Table 3). None of the participants had serious adverse events during and immediately after the PTPs. There was one Ex-G2 participant who died of cardiac arrest on the following day after the first bedside PTPs. There were no physiotherapists who tested positive for COVID-19 as a consequence of instructing bedside PTPs to patients. The number of physiotherapy sessions was significantly different between the groups (*p* < 0.001).

## Discussion

To our knowledge, this is the first study to investigate the different frequencies of bedside PTPs in COVID-19 patients during admission to the hospital. In addition, there was a lack of study reports on the safety of bedside PTPs in the acute phase of COVID-19 patients who were mostly identified with a mild to moderate degree of severity. The main findings found no differences between groups in regard to survival rate, LoH, referrals to the ICU, and in-hospital complications. Overall, there was a high survival rate of patients, no deaths among mild to moderate COVID-19 patients, a limited number of COVID-19 patients who were referred to ICU after receiving the PTPs, and a low rate of complications observed after receiving the PTPs. Importantly, none of the serious adverse events occurred during and immediately after each PTP session. In addition, no physiotherapists tested positive for COVID-19 during two months of the in-hospital data collection period.

To date, there are few reports on the effects of PTPs documented in the acute phase of COVID-19 [11, 12, 14]. In literature, most previous studies have investigated physical rehabilitation in severe cases of COVID-19 or in patients who were referred to ICU[11, 14] or after recovery from critical illnesses [20, 21]. Also, a study assessed patients who were negative for SAR-CoV-2 by laboratory diagnostic tests [21] or long-lasting hospital stay, which was safer and less likely to be infected with COVID-19 compared to our study conducted with active COVID-19. Interestingly, previous studies have reported that PTPs were safe and feasible in the ICU setting or the post-recovery period [14, 20, 21]. PTPs also improved the patient’s motor and respiratory function, along with functional activity, particularly in post-critical illness patients [14, 20, 21]. However, those patients were older and had more severe conditions than patients in the present study [11, 12, 14].

The effects of PTPs in the current study may provide positive effects on physical function similar to a previous study that had examined a one-week telerehabilitation in mild to moderate COVID-19 patients who were confined at home [25]. To support this, there was only one patient who was referred to the ICU, few complications found, and none of the patients with mild to moderate conditions died in the present study. In addition, our study showed minor complications (9.6%, 4 bacterial infections, 1 influenza) compared to an earlier study that had found approximately 39% of complications in patients aged between 19 and 49 years [26]. Also, there were no patients who developed ARDS in this study, which is contrary to a previous study that had reported 23% and 4% of ARDS in the pneumonia group and mild to moderate group, respectively [27]. For these reasons, the survival rate of patients with mild to moderate conditions in the current study was very high, contrary to many previous studies [12, 14]. Meanwhile, the LoH of a previous study [20] is comparable to the present study, which was 9.8 days. Similarly, the median LoH of patients with pneumonia not caused by COVID-19 was 9 days [28]. In contrast, a recent review study had found that the median LoH of COVID-19 patients in China and other countries was 14 and 5 days, respectively. However, the LoH during the first year of the COVID-19 pandemic showed higher LoH compared to the current study [29]. The variations in LoH could be due to differences in health policies for each country, the development of treatment, and the effectiveness of vaccination. In Thailand, the Ministry of Public Health has announced that COVID-19 patients must stay in the hospital for at least 10 days as a control against SAR-CoV-2 transmission.

The two most used PTPs including breathing exercises and progressive mobilization/ exercise confirmed that they were safe for COVID-19 patients. None of the patients experienced serious adverse events during and immediately after the PTPs. This might be caused by the comprehensive screening of patients’ clinical records before and after the PTPs [20]. Among the minor adverse events, a drop of SpO_2_ >3% from baseline was the most common adverse effect found in the Ex-G2 group because participants in the Ex-G2 group were encouraged at the bedside to perform light exercise/ ambulation. Our findings support a recent systemic review that had found that pulmonary rehabilitation was safe and feasible for COVID-19 patients [30]. In addition, it is in line with a previous recommendation that patients with mild conditions may benefit from breathing exercises [31]. A previous study has also shown symptomatic improvement after six weeks of an online breathing program for patients with post-COVID-19 condition [32]. In addition, patients with mild symptoms of SARS-CoV-2 were able to perform mild-to-moderate intensity exercise during active COVID-19 [33]. However, the exercise intensity which is higher than the lactate threshold might not be appropriate according to the production of the respiratory droplet [34]. Also, previous studies have demonstrated that exercise at moderate intensity could downregulate inflammatory cytokines and stimulate the release of anti-inflammatory substances [18, 19]. Likewise, two weeks of moderate aerobic exercise for COVID-19 patients recently have shown an improvement in immune function [17]. A recent study supported that exercise promotes myokines production which would alleviate SARS-CoV-2 infectivity [35]. Besides the physical aspect, the psychological impact may also improve after bedside PTPs which is supported by a previous study [36]. The PTPs in this study are similar to our study, except for craft activities, which were added in the previous study [36]. Thus, a positive immune system response, the release of anti-SAR CoV-2 substances, and mental improvement after the PTPs might promote the beneficial effects on most of the primary outcome measures. Notably, we provided the BreatheMax, a breathing biofeedback device, to Ex-G2 patients. From our observation, it can increase SpO_2_ within a few minutes, similar to a conventional breathing exercise. Thus, this study supports a previous recommendation that PEP or OIS devices can be applied to COVID-19 patients without complications [9]. Interestingly, one to two times of bedside PTPs provided equal benefits compared to the daily PTP group. This may be caused by the patients of both groups being encouraged to perform PTPs via a ward phone, private mobile phone application, and closed private groups. Thus, daily bedside PTPs may be unnecessary to implement for COVID-19 patients with mild to moderate conditions. Telemedicine is an alternative platform in the case of restrictions for bedside PTPs. Importantly, our study confirmed that bedside PTPs were safe for physiotherapists because there were no reports of COVID-19 infection during the two months of prospective data collection among these physiotherapists. Indeed, the physiotherapists in our study strictly followed the airborne, aerosol, and manual contact precautions when providing bedside PTPs to reduce the possibility of SARS-CoV-2 infection.

There were some limitations in the present study. First, this study did not have a control group, and thus there were no data to compare with conventional treatment. Second, the objective outcome measures and psychological status at baseline could not be performed due to unstable signs and symptoms and concerns about COVID-19 transmission, which were similar to most previous case reports that were unable to perform those tests [11, 12]. In addition, blood biomarkers and chest radiographs were not allowed to be tested before discharge according to the limitation of hospital rules. These variables may have been useful to explain the physiological changes after the PTPs. Third, according to the quarantine rule, patients could not be discharged from the hospital even if there were no complications and they had almost fully recovered from COVID-19 infection. Consequently, the LoH among the groups found no differences. Future studies are highly recommended to confirm the findings of the present study with a conventional treatment control group and regardless of a mandatory quarantine rule.

## Conclusion

The different bedside PTP frequency in COVID-19 patients with primarily mild to moderate conditions found no differences in the survival rate, LoH, referrals to the ICU, and in-hospital complications. PTPs are safe for COVID-19 patients and physiotherapists. A prudent assessment and monitoring of physiological parameters during the PTPs are necessary to prevent unpredictable adverse events.

## Data Availability

All relevant data are within the manuscript and its Supporting Information files.

## Supporting information

S1 raw data

(XLSX)

S2 Trend checklist

(PDF)

## Acknowledgments

We would like to thank all the researchers who participated in this study, in particular, the physiotherapists involved in the data collection processes. They were willing to join the study even though they might have a chance to contract COVID-19. The authors are thankful to the Bamrasnaradura Infectious Disease Institute, which allowed us to collect data. Importantly, we are indebted to all COVID-19 patients, who devoted time to participate in this study.

## Author Contributions

**Conceptualization:** Khomkrip Longlalerng, Netchanok Jianramas, Veeranoot Nissapatorn, Chaisith Sivakorn, Maria de Lourdes Pereira, Chenpak Salesingh, Eittipad Jaiyen.

**Data curation:** Khomkrip Longlalerng, Netchanok Jianramas, Anuttra (Chaovavanich) Ratnarathon, Chenpak Salesingh, Eittipad Jaiyen, Salinee Chaiyakul, Nitita Piya-amornphan, Thanaporn Semphuet, Thanrada Thiangtham, Kornchanok Boontam.

**Formal analysis:** Khomkrip Longlalerng, Netchanok Jianramas.

**Funding acquisition:** Khomkrip Longlalerng, Maria de Lourdes Pereira.

**Investigation:** Khomkrip Longlalerng, Netchanok Jianramas, Anuttra (Chaovavanich) Ratnarathon, Chenpak Salesingh, Eittipad Jaiyen, Salinee Chaiyakul, Nitita Piya-amornphan, Thanaporn Semphuet, Thanrada Thiangtham, Kornchanok Boontam.

**Methodology:** Khomkrip Longlalerng, Netchanok Jianramas, Veeranoot Nissapatorn, Chaisith Sivakorn, Maria de Lourdes Pereira, Anuttra (Chaovavanich) Ratnarathon, Chenpak Salesingh, Eittipad Jaiyen, Salinee Chaiyakul, Nitita Piya-amornphan, Thanaporn Semphuet, Thanrada Thiangtham.

**Project administration:** Khomkrip Longlalerng, Netchanok Jianramas.

**Writing – original draft:** Khomkrip Longlalerng.

**Writing – review & editing:** Khomkrip Longlalerng, Netchanok Jianramas, Veeranoot Nissapatorn, Chaisith Sivakorn, Maria de Lourdes Pereira, Anuttra (Chaovavanich) Ratnarathon, Chenpak Salesingh, Eittipad Jaiyen, Salinee Chaiyakul, Nitita Piya-amornphan, Thanaporn Semphuet, Thanrada Thiangtham, Kornchanok Boontam.

